# Deep Learning in the prediction of Angiography-proven Severe Coronary Stenosis in Patients with Apparently Normal Electrocardiograms

**DOI:** 10.1101/2022.11.26.22282701

**Authors:** Zhengkai Xue, Shijia Geng, Shaohua Guo, Guanyu Mu, Bo Yu, Peng Wang, Sutao Hu, Weilun Xu, Yanhong Liu, Lei Yang, Huayue Tao, Kangyin Chen, Shenda Hong

## Abstract

Patients with severe coronary artery stenosis may have apparently normal electrocardiograms (ECGs), making it difficult to detect the adverse health conditions during screening or physical examinations, resulting in them missing the optimal window of treatment. The goal of this study was to develop an artificial intelligence-based ECG model which can distinguish severe coronary stenosis (≥ 90%) from no or mild coronary stenosis (< 50%) in patients with apparently normal ECGs. Deep learning (DL) models trained from scratch with pre-trained parameters (transfer learning) were tested on ECG alone as well as on ECG along with clinical information (age, sex, hypertension, diabetes, dyslipidemia and smoking status). We also compared the performance of logistic regression for clinical information only and found that DL models trained from scratch with ECG alone can achieve a specificity of 0.746; however, they have low sensitivity, which is comparable to the performance of logistic regression with clinical data. Although adding clinical information to the ECG DL model trained from scratch can improve the sensitivity, it can reduce the specificity. Combining clinical information with the ECG transfer learning model provides the best performance, with a 0.847 AUC, 0.848 sensitivity, and 0.704 specificity.

## 1 Introduction

Coronary artery disease (CAD) is one of the most common cardiovascular diseases in the world and can cause severe life-threatening cardiovascular events such as acute myocardial infarction (MI), malignant arrhythmia, and heart failure [1, 2]. The timely treatment of CAD can improve the prognosis and reduce the incidence of adverse events and mortality. Given this, the early screening and diagnosis of CAD are of great significance. Coronary angiography (CAG) is the “gold standard” for the diagnosis of CAD; however, it cannot be used for screening since it is invasive, expensive and requires specialized equipment and personnel. Electrocardiogram (ECG), on the other hand, is a simple and easy test to perform and is widely used for the screening and diagnosis of heart diseases. However, ECG has low sensitivity and specificity in predicting coronary artery stenosis [3], especially in CAD patients without myocardial ischemia, which may appear normal or apparently normal (when neither the ECG machine’s automatic interpretation nor the doctor’s manual interpretation report significant abnormalities). Figure 1 shows two patients with apparently normal resting ECGs. Patient A’s CAG images show no obvious coronary lesions; however, Patient B’s CAG images reveal severe coronary stenosis, with about 95% stenosis in the distal part of the circumflex branch and about 80% stenosis in the middle part of the left anterior descending branch. In clinical practice, it is common for CAD patients with severe coronary stenosis to have only mild angina and apparently normal ECGs. This situation is extremely harmful because the patients’ conditions are not easily detected during routine physical examinations or screenings, and the best time for treatment may have been missed by the time severe symptoms appear.

**Figure 1:**
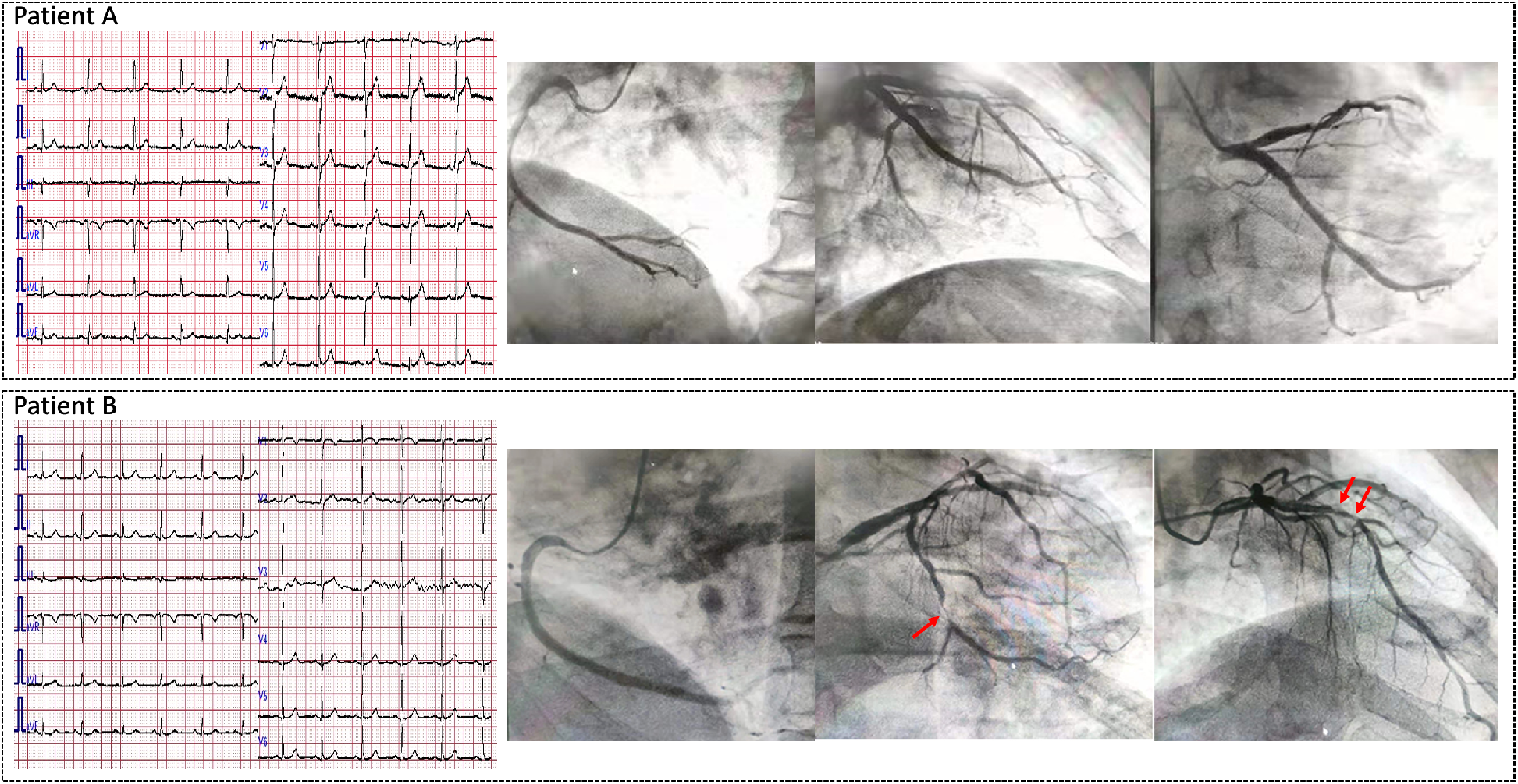
ECGs and CAG Images of Two Patients. Both patients have an apparently normal ECG. Patient A’s CAG images show no obvious coronary lesions; however, Patient B’s CAG images show severe coronary stenosis in both the circumflex branch and the left anterior descending branch (see red arrows).

In recent years, methods using artificial intelligence (AI) have been developed for use in the field of cardiovascular studies [4], in which an ECG model based on deep learning (DL) is used in the diagnosis of arrhythmia [5, 6], cardiac systolic or diastolic dysfunction [7, 8], valvular heart disease [9], cardiomyopathy [10], predicting the risk of death [11], and so on [12]. For CAD, Leasure’s group developed a DL model to correlate ECGs with the results of CAG, to classify the severity and location of coronary artery stenosis [13]. Also, Huang and colleagues tested six well-known computer vision DL models to predict the location of obstructed coronary artery from ECG images [14]. The models developed by both groups obtained high sensitivity and high specificity. However, these studies do not emphasize ECG status and might include ECGs that show obvious abnormalities. The abnormalities could ease the model training and make the AI-model less powerful since doctors can find the problems by themselves without AI assistant.

We carefully select apparently normal ECGs for our study with the aim of exploring the performances of DL models in identifying patients with severe (≥ 90%) stenosis versus no/mild (< 50%) stenosis. Our findings are as follows

1. DL models have the potential to predict patients with severe coronary stenosis with apparently normal ECGs.
2. DL model trained from scratch with ECG alone can achieve a specificity of 0.746; however, the sensitivity is low. The performance is similar to that of logistic regression of clinical information including age, gender, hypertension, diabetes, dyslipidemia, and smoking status.
3. Adding clinical information to the ECG DL model trained from scratch can improve the sensitivity; however, it drastically reduces the specificity.
4. Combining clinical information with the outputs of an existing ECG classification model (transfer learning) can achieve an accuracy of 0.750 with a sensitivity of 0.848 and specificity of 0.704 for identifying severe coronary artery stenosis.

## 2 Materials and Methods

### 2.1 Data sources

Patients admitted to the Department of Cardiology at the Second Hospital of Tianjin Medical University from January 2019 to February 2021, who received at least one standard 12-lead ECG within 3 days before CAG were included in the study. The selection criteria were as follows: over 18 years of age; apparently normal ECG; no acute or old MI; no previous percutaneous coronary intervention (PCI) or coronary artery bypass grafting (CABG). We also collect data on age, gender, past medical history, smoking status, and results from laboratory testing. All clinical data were obtained with the informed consent of patients and approved by the hospital ethics committee. The summary of the clinical baseline characteristics for the whole dataset is shown in Table 1.

**Table 1:** Baseline Characteristics for All Data

| Baseline Characteristics | Total<br>(n=392) | Case<br>(n=138) | Control<br>(n=254) | P value |
| --- | --- | --- | --- | --- |
| Age (years old) | 62<br>(56-67) | 65<br>(60-69) | 60<br>(53-65) | <0.001* |
| # of Males | 163<br>(41.6%) | 80<br>(58.0%) | 83<br>(32.7%) | <0.001* |
| # of Patients with Hypertension | 246<br>(62.8%) | 109<br>(79.0%) | 137<br>(53.9%) | <0.001* |
| # of Patients with Diabetes | 88<br>(22.4%) | 56<br>(40.6%) | 32<br>(12.6%) | <0.001* |
| # of Patients with Dyslipidemia | 159<br>(40.6%) | 98<br>(71.0%) | 61<br>(24.0%) | <0.001* |
| # of Patients Who Smoke | 96<br>(24.5%) | 38<br>(27.5%) | 58<br>(22.8%) | 0.301 |

A total of 392 patients were selected for the study, and each patient had a 10-second standard 12-lead ECG that was acquired using a Fukuda Denshi CardiMax FX-7402 ECG Machine. A professional cardiologist qualified for ECG diagnosis manually interpreted the ECG graphs printed out with 25mm/s paper speed and 10mm/mV voltage calibration. We decrypted and extracted raw ECG data that had a 500Hz sampling frequency using the EFS-200C software provided by the manufacturer. The summary of the ECG measurements is listed in Table 2.

**Table 2:** ECG Measurements for All Data (*±* is for verified normally distributed data, (*−*) is the interquartile range for non-normally distributed data)

| ECG Measurements | Total<br>(n=392) | Case<br>(n=138) | Control<br>(n=254) | P value |
| --- | --- | --- | --- | --- |
| HR (bpm) | 69<br>(63-76) | 70<br>(64-78) | 68<br>(62-75) | 0.061 |
| RR (ms) | 864 ± 114 | 849 ± 119 | 873 ± 111 | 0.053 |
| PR (ms) | 161<br>(148-178) | 166<br>(149-183) | 160<br>(146-175) | 0.017* |
| QRS (ms) | 97<br>(92-104) | 96<br>(91-103) | 97<br>(92-104) | 0.284 |
| QT (ms) | 397 ± 27 | 394 ± 26 | 398 ± 27 | 0.222 |
| QTc (ms) | 428 ± 20 | 429<br>(415-445) | 428<br>(414-441) | 0.419 |
| Axis (degree) | 35<br>(9-56) | 29<br>(0-52) | 37<br>(13-59) | 0.015* |
| SV1 (mV) | 0.59<br>(0.42-0.85) | 0.63<br>(0.43-0.86) | 0.58<br>(0.42-0.85) | 0.462 |
| RV5 (mV) | 1.33<br>(1.03-1.62) | 1.39<br>(1.09-1.63) | 1.26<br>(1.01-1.60) | 0.074 |
| RV5 + SV1 (mV) | 1.95<br>(1.60-2.34) | 2.08<br>(1.69-2.38) | 1.91<br>(1.55-2.32) | 0.051 |

Based on the CAG results, we classified coronary artery stenosis as severe (case group) and no/mild (control group) stenosis by visual inspection of lumen diameter. Severe stenosis was defined as ≥ 90% stenosis in at least one of the major coronary arteries or their major branches, and no/mild stenosis as < 50% stenosis in all major coronary arteries and their major branches. The CAG results were independently assessed and reported by two professional interventional cardiologists during the process with reference to computer-assisted quantitative coronary angiography (QCA) results, and the final degree of stenosis was determined with the agreement of the two physicians. The lesion information from CAG is shown in Table 3. Under the guidance of cardiologists, technicians labelled the ECGs with corresponding labels (0 for no/mild stenosis, and 1 for severe stenosis).

**Table 3:** Lesion Information from CAG for All Data

| Lesion | Number | Percentage |
| --- | --- | --- |
| 1 vessel | 33 | 23.9% |
| LAD | 25 | 18.1% |
| LCX | 4 | 2.9% |
| RCA | 4 | 2.9% |
| 2 Vessels | 31 | 22.5% |
| 3 Vessels | 59 | 42.8% |
| Ramus | 1 | 0.7% |
| Ramus + 1 Vessel | 1 | 0.7% |
| Ramus + 2 Vessels | 1 | 0.7% |
| Ramus + 3 Vessels | 1 | 0.7% |
| LM + 1 Vessel | 1 | 0.7% |
| LM + 2 Vessels | 4 | 2.9% |
| LM + 3 Vessels | 6 | 4.4% |

We randomly assigned the data into training and test sets in a ratio of approximately 3:1, and obtained 288 training examples and 104 test examples. The flow diagram for the overall data selection process is shown in Figure 2, and Table 4 lists the baseline characteristics for the training and test set, respectively.

**Table 4:** Baseline Characteristics for Training and Test Set

| Baseline Characteristics | Training Set<br>(N=288) | Test Set<br>(N=104) |
| --- | --- | --- |
| # of Cases | 105<br>(36.5%) | 33<br>(31.7%) |
| Age (years old) | 61<br>(56-67) | 62<br>(55-66) |
| # of Males | 122<br>(42.4%) | 41<br>(39.4%) |
| # of Patients with Hypertension | 180<br>(62.5%) | 66<br>(63.5%) |
| # of Patients with Diabetes | 66<br>(22.9%) | 22<br>(21.2%) |
| # of Patients with Dyslipidemia | 113<br>(39.2%) | 46<br>(44.2%) |
| # of Patients who Smoke | 71<br>(24.7%) | 25<br>(24.0%) |

**Figure 2:**
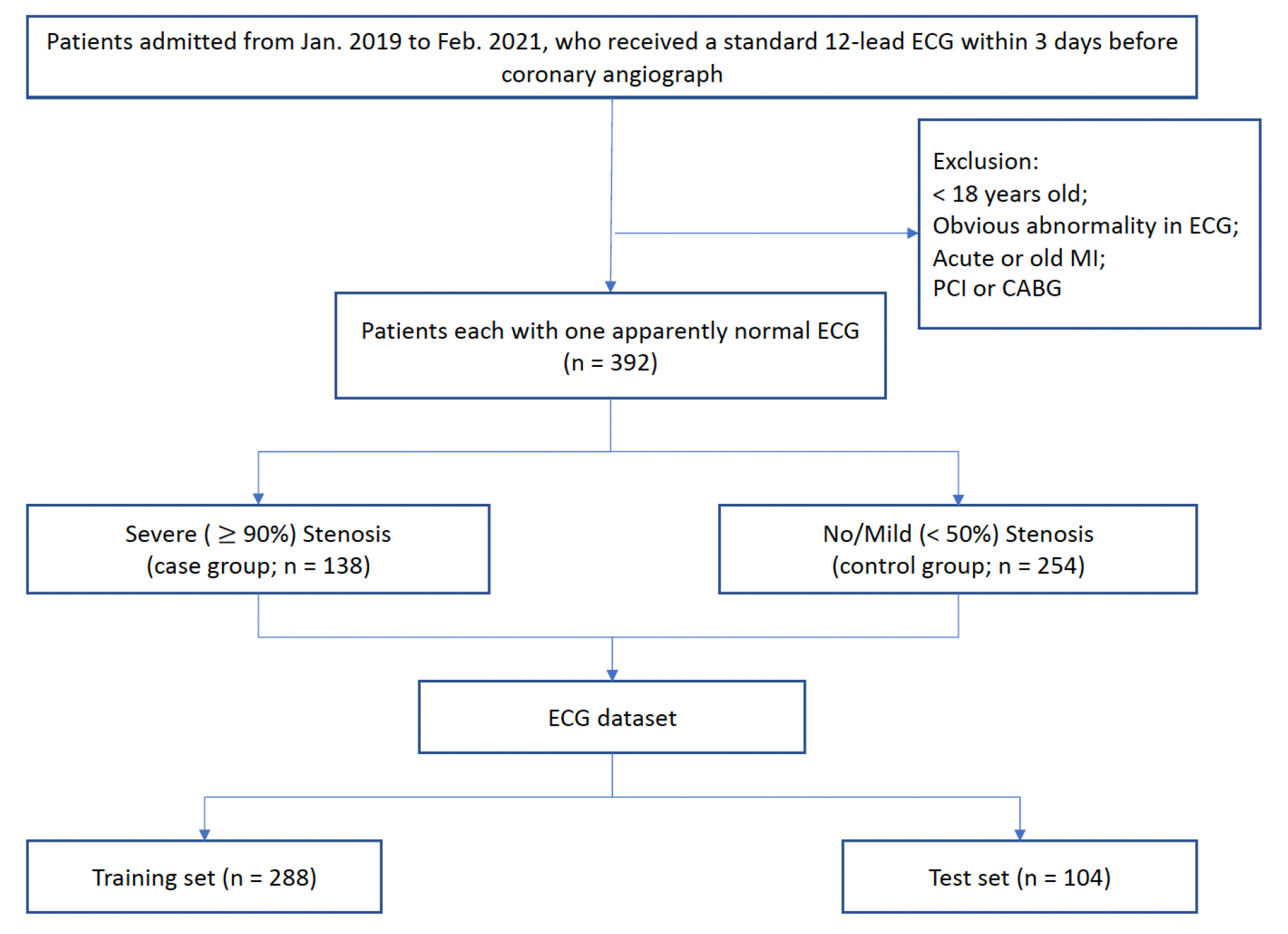
Flow Diagram of the Criteria

### 2.2 Models

In the study, we conducted experiments to differentiate the severity of coronary stenosis using ECGs based on a deep convolutional neuron network (CNN) structure verified to perform well on ECG-related tasks [15, 16, 17]. The input layer of the model structure contained 12 channels representing the 12 leads of the ECG, and each channel had 5000 data points that matched a 10-second recording with a sampling frequency of 500 Hz. The original output layer had 47 units corresponding to 47 ECG classes in the original classification task. To perform our task, we added a fully connected layer at the end with one output unit that produces the probability of severe coronary stenosis (1 - probability of no/mild coronary stenosis). With different inputs and training methods, we obtained four models: a model trained from scratch with ECG alone; a model trained from scratch with ECG and baseline features; a transfer learning model with ECG alone; and a transfer learning model with ECG and baseline features. Models training from scratch had random weights at the beginning and used the training data to optimize the cost function, which is the binary cross-entropy between the predicted stenosis probabilities and the actual labels. Transfer learning models borrowed the weights pre-trained from the ECG classification task, which presumably contained information on how to process and analyze ECG data, and further optimized the cost function using the training data based on these weights. We call the 47 features extracted from the original deep CNN model as “deep features”, and the details of the features are discussed in Section 4. For the models with additional baseline features, we added the six baseline characteristics of patients (age (normalized), gender (0 female; 1 male), hypertension (0 no; 1 yes), diabetes (0 no; 1 yes), dyslipidemia (0 no; 1 yes), smoking status (0 no; 1 yes)) on the second last layer to combine them with the 47 deep features, and fed the total of 53 features into the last fully-connected layer to obtain the final decision.

We also conducted logistic regression on the baseline features alone to study the effects of baseline characteristics on severe coronary stenosis differentiation and verified the outcomes from the combination of deep and baseline features. All of the five experimental models are illustrated in Figure 3.

**Figure 3:**
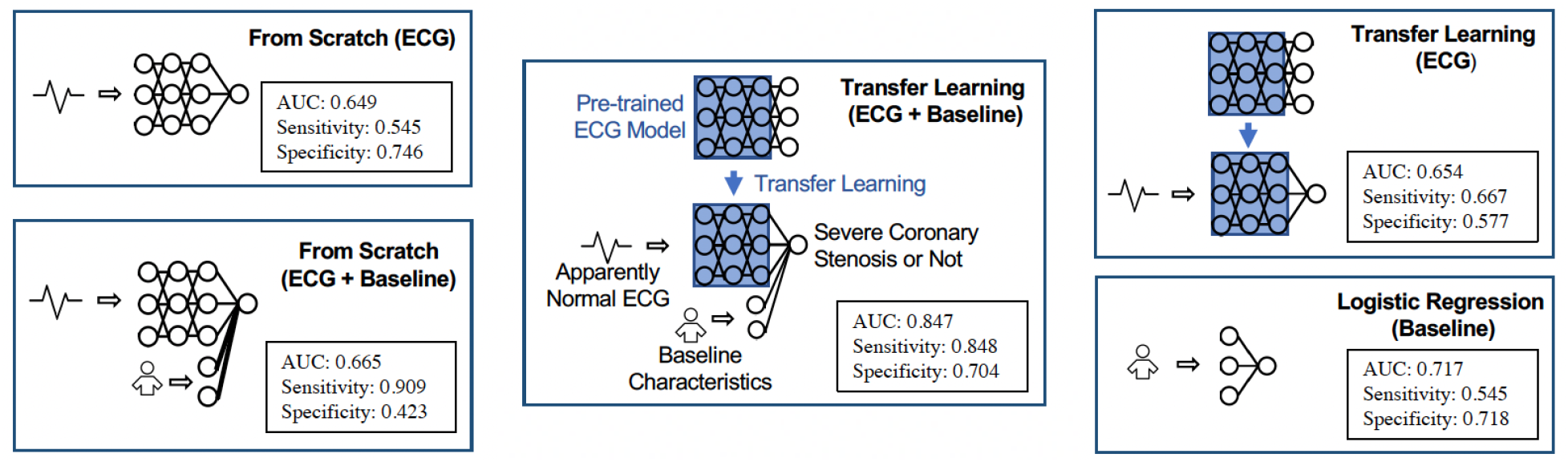
Diagram of Experimental Models

### 2.3 Evaluation Metrics

After training, we calculated and reported the results metrics on the test data set, which included area under the curve (AUC), accuracy, precision, sensitivity, specificity, and F1-score. The AUC refers specifically to the area under the receiver operating characteristics (ROC) curve. A ROC curve plots the performance of a binary classification model at different threshold settings, and AUC represents the overall classification ability of the model. The x-axis of the ROC curve is the false positive rate (FPR), and the y-axis is the true positive rate (TPR). In our study, the false positive (FP) is the actual no/mild stenosis that is predicted as severe stenosis by the model, and the true positive (TP) is the number of actual severe stenosis that is predicted as severe stenosis. FPR is the ratio of FP to the total actual no/mild stenosis, which is the sum of FP and the true negative (TN), while TPR is the ratio of TP to the total actual severe stenosis, which is the sum of TP and the false negative (FN). We preferred a higher TPR as well as a lower FPR and selected a threshold that maximized the Youden’s Index (TPR - FPR). Based on this selected threshold, we calculated and reported the accuracy, precision, sensitivity, specificity, and F1-score. The equations used for the calculations are as follows:

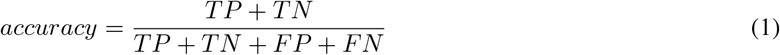

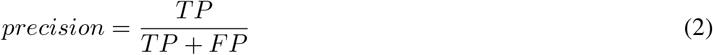

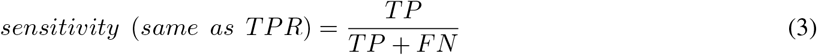

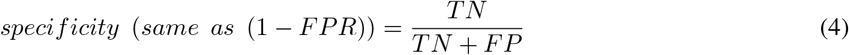

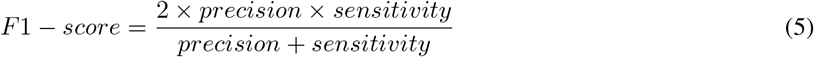

## 3 Results

With the testing data, we obtained the probabilities of severe coronary stenosis for each model. Figure 4 shows the ROC curves of all the models. The ECG and baseline features model with transfer learning gave the highest AUC (0.847), while other DL models had similar AUC (0.649 - 0.665). The logistic regression model for baseline features had an AUC of 0.717, which was the second highest value.

**Figure 4:**
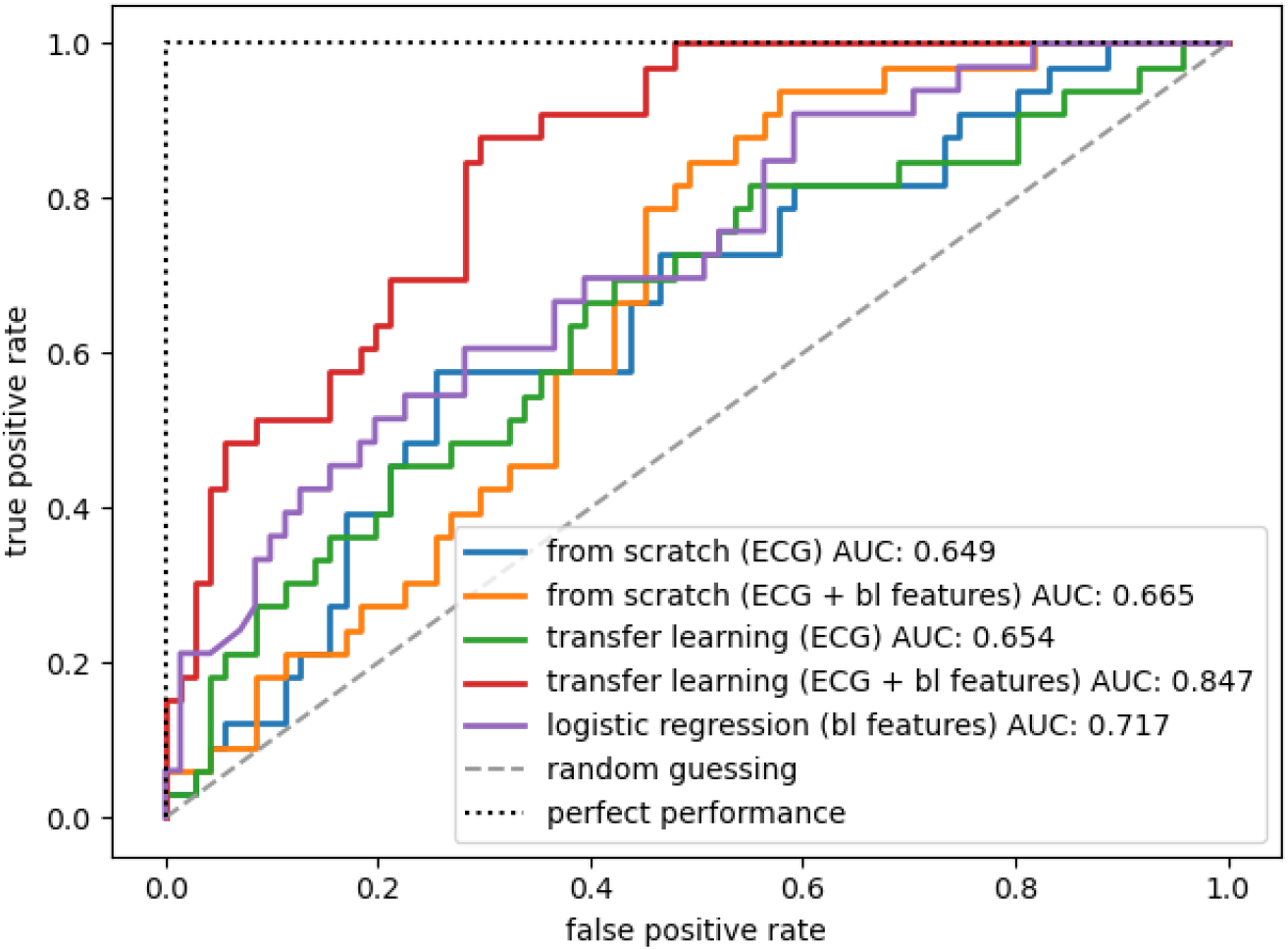
ROC Curves of All Models

Table 5 shows the metrics of the test results for each model with the optimal threshold selected based on Youden Index. The ECG and baseline features model with transfer learning gave the best accuracy (0.750), precision (0.571) and F1-score (0.683) among the experimental models. The ECG and baseline features model trained from scratch had the highest sensitivity (0.909), while the ECG-only model trained from scratch had the highest specificity (0.746). The AUC, sensitivity, and specificity for each model are shown in Figure 3, and the normalized confusion matrices of the models based on the selected thresholds are shown in Figure 5.

**Table 5:** Metrics of All Models (*±*95% CI)

| Model | Threshold | Accuracy | Precision | Sensitivity | Specificity | F1 Score |
| --- | --- | --- | --- | --- | --- | --- |
| from scratch (ECG) | 0.241 | 0.683 ± 0.089 | 0.500 ± 0.163 | 0.545 ± 0.170 | **0.746** ± 0.101 | 0.522 ± 0.167 |
| from scratch (ECG + bl) | 0.299 | 0.577 ± 0.095 | 0.423 ± 0.115 | **0.909** ± 0.098 | 0.423 ± 0.115 | 0.577 ± 0.134 |
| transfer learning (ECG) | 0.299 | 0.606 ± 0.094 | 0.423 ± 0.134 | 0.667 ± 0.161 | 0.577 ± 0.115 | 0.518 ± 0.150 |
| transfer learning (ECG + bl) | 0.345 | **0.750** ± 0.083 | **0.571** ± 0.139 | 0.848 ± 0.122 | 0.704 ± 0.106 | **0.683** ± 0.142 |
| logistic regression (bl) | 0.597 | 0.663 ± 0.091 | 0.474 ± 0.159 | 0.545 ± 0.170 | 0.718 ± 0.105 | 0.507 ± 0.164 |

**Figure 5:**
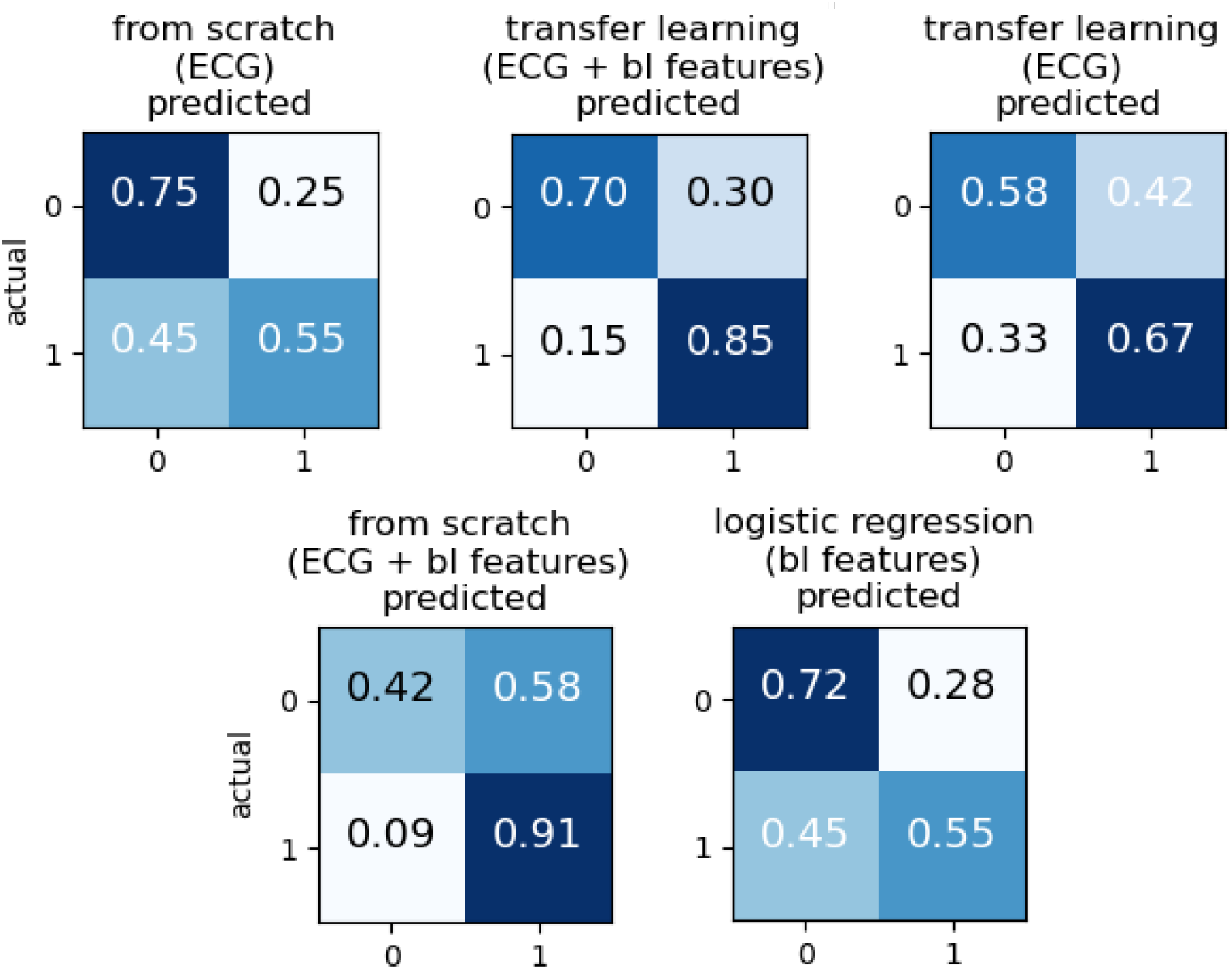
Normalized Confusion Matrices of All Models

We also conducted regression analysis on the baseline features and have presented theses results in Table 6. Age, hypertension and dyslipdemia have significant coefficients among all the baseline features.

**Table 6:** Regression Analysis Results for Baseline Features

|  | coef | std err | z | P > \|z\| | [0.025 | 0.975] |
| --- | --- | --- | --- | --- | --- | --- |
| age | 0.8208 | 0.159 | 5.159 | 0.000* | 0.509 | 1.133 |
| genders | 0.1490 | 0.296 | 0.504 | 0.614 | -0.431 | 0.729 |
| hypertension | -0.8802 | 0.227 | -3.877 | 0.000* | -1.325 | -0.435 |
| diabetes | 0.6524 | 0.335 | 1.949 | 0.051 | -0.004 | 1.308 |
| dyslipidemia | 1.0405 | 0.274 | 3.804 | 0.000* | 0.504 | 1.577 |
| smoke | -0.3021 | 0.340 | -0.889 | 0.374 | -0.968 | 0.364 |

## 4 Discussion

In the study, we investigated the feasibility of using DL techniques to distinguish between severe (≥90%) and no/mild (< 50%) coronary stenosis from apparently normal ECG. We discovered that with ECG alone, the DL model trained from random parameters can achieve a specificity of 0.746, the highest among all tested models. This suggests that the DL model trained from scratch can accurately identity no/mild stenosis conditions in the control group using ECG alone. However, its sensitivity was only 0.545, indicating that it might not perform well in predicting severe stenosis in the case group. We hypothesize that adding more features can improve the ability to identify severe cases, and by adding patients’ baseline characteristics (age, gender, hypertension, diabetes, dyslipidemia and smoking status), we greatly increased the sensitivity to 0.909. However, the specificity dropped to 0.423, which is the lowest among the experimental models. This might be due to the lack of training data, which causes a high bias for the prediction model.

Therefore, we applied the transfer learning technology and fine-tuned the model based on parameters from an ECG classification task. With ECG-alone transfer learning model, we obtained an acceptable sensitivity of 0.667; however, the specificity was still relatively low (0.577). Finally, we tried the ECG and baseline features transfer learning model and obtained a relatively high specificity (0.704) as well as a a high sensitivity (0.848).

By comparison, we also applied logistic regression on the baseline features alone and obtained an accuracy of 0.663 with a sensitivity of 0.545 and sensitivity of 0.718. The baseline characteristics selected (age, gender, hypertension, diabetes, dyslipdemia, smoking status) are generally accepted cardiovascular risk factors [18]. In Table 6, we have presented the analysis results for the risk factors from the logistic regression. Age, hypertension, and dyslipidemia had significant coefficients, which indicates that these three factors influence the discrimination of coronary stenosis severity. The logistic regression with baseline features had similar performances as that of the ECG-alone model trained from scratch, which was better than the ECG-alone transfer learning model. The reason may be that the ECG classification task and the coronary stenosis discrimination task are essentially different, and the model pre-trained for the ECC classification task may not capture the ECG characteristics that determine the severity of the stenosis. However, by adding the baseline features to the transfer learning model, we can correct the skewed information and achieve the highest performance among the experimental models.

Similar to the results of logistic regression analysis for baseline features, we also report the results of deep features regression analysis in Table 7. We observed that among the deep features, PVC (Premature Ventricular Contraction), VT (Ventricular Tachycardia), AFL (Atrial Flutter), VE (Ventricular Escape), LVH (Left Ventricular Hypertrophy), RAH (Right Atrial Hypertrophy), PACED (Paced Rhythm), and PAUSE (Sinus Pause) have significant coefficients. It can be interpreted that these deep features are effective in identifying severe coronary stenosis. However, although corresponding to ECG classes, the deep features are intermediate features that cannot represent the diagnosis decisions on certain classes. Further research is required to explore the relationship between the pre-trained classification task and the stenosis discrimination task. For the time being, the ECG classes should only be treated as tags for the deep features and should not be overanalyzed.

**Table 7:** Regression Analysis Results for Deep and Baseline Features

|  | coef | std err | z | P> \|z\| | [0.025 | 0.975] |
| --- | --- | --- | --- | --- | --- | --- |
| N | 1.2033 | 5.080 | 0.237 | 0.813 | -8.752 | 11.159 |
| SN | -12.6390 | 14.156 | -0.893 | 0.372 | -40.384 | 15.106 |
| SNA | -8.9506 | 7.403 | -1.209 | 0.227 | -23.461 | 5.560 |
| SNT | -6.8657 | 9.297 | -0.739 | 0.460 | -25.087 | 11.355 |
| SNB | -5.6310 | 4.154 | -1.356 | 0.175 | -13.773 | 2.511 |
| PVC | -9.0575 | 4.566 | -1.983 | 0.047* | -18.008 | -0.107 |
| PSC | 21.5940 | 22.342 | 0.967 | 0.334 | -22.196 | 65.384 |
| PJC | -28.9159 | 16.561 | -1.746 | 0.081 | -61.375 | 3.543 |
| PAC | -9.8879 | 21.681 | -0.456 | 0.648 | -52.382 | 32.606 |
| VT | 16.0586 | 7.891 | 2.035 | 0.042* | 0.593 | 31.525 |
| SVT | -11.4579 | 10.078 | -1.137 | 0.256 | -31.210 | 8.294 |
| AFL | -42.8928 | 13.144 | -3.263 | 0.001* | -68.655 | -17.130 |
| AF | 3.2439 | 5.424 | 0.598 | 0.550 | -7.387 | 13.875 |
| VFL | 41.2271 | 22.961 | 1.796 | 0.073 | -3.776 | 86.230 |
| VF | -31.1411 | 19.798 | -1.573 | 0.116 | -69.944 | 7.662 |
| WPW | -1.2871 | 4.740 | -0.272 | 0.786 | -10.578 | 8.003 |
| VE | 25.8193 | 10.904 | 2.368 | 0.018* | 4.448 | 47.191 |
| SE | -8.3346 | 22.024 | -0.378 | 0.705 | -51.501 | 34.832 |
| JE | 15.8398 | 18.665 | 0.849 | 0.396 | -20.743 | 52.422 |
| AE | 23.9337 | 19.197 | 1.247 | 0.212 | -13.691 | 61.558 |
| AVBI | 1.4219 | 12.812 | 0.111 | 0.912 | -23.690 | 26.533 |
| AVBII | 17.2381 | 14.014 | 1.230 | 0.219 | -10.229 | 44.705 |
| AVBIII | -24.4169 | 18.452 | -1.323 | 0.186 | -60.583 | 11.749 |
| IVB | 5.1382 | 3.192 | 1.610 | 0.107 | -1.119 | 11.395 |
| LBBB | -6.3391 | 7.323 | -0.866 | 0.387 | -20.692 | 8.014 |
| RBBB | -1.4731 | 3.709 | -0.397 | 0.691 | -8.742 | 5.796 |
| LAFB | -2.9586 | 3.145 | -0.941 | 0.347 | -9.123 | 3.206 |
| LVH | 10.3416 | 4.964 | 2.083 | 0.037* | 0.612 | 20.071 |
| RVH | 1.9239 | 3.711 | 0.518 | 0.604 | -5.349 | 9.196 |
| LAH | 6.4768 | 5.535 | 1.170 | 0.242 | -4.372 | 17.325 |
| RAH | 17.2804 | 8.001 | 2.160 | 0.031* | 1.598 | 32.963 |
| ST | 7.7787 | 7.165 | 1.086 | 0.278 | -6.265 | 21.822 |
| T | 9.9505 | 6.797 | 1.464 | 0.143 | -3.371 | 23.272 |
| Q | -15.2213 | 10.697 | -1.423 | 0.155 | -36.188 | 5.745 |
| MI | 16.5157 | 11.427 | 1.445 | 0.148 | -5.880 | 38.912 |
| STTQ | -18.3151 | 13.891 | -1.319 | 0.187 | -45.540 | 8.910 |
| PACED | 17.1037 | 6.730 | 2.542 | 0.011* | 3.914 | 30.293 |
| LAD | -3.3198 | 2.353 | -1.411 | 0.158 | -7.932 | 1.293 |
| RAD | -4.5820 | 3.706 | -1.236 | 0.216 | -11.845 | 2.681 |
| CW | 5.0501 | 3.568 | 1.415 | 0.157 | -1.944 | 12.044 |
| CCW | 0.9500 | 3.289 | 0.289 | 0.773 | -5.497 | 7.397 |
| BLV | 5.3387 | 5.501 | 0.971 | 0.332 | -5.443 | 16.120 |
| CLV | 1.9716 | 5.572 | 0.354 | 0.723 | -8.948 | 12.892 |
| PAUSE | -35.3013 | 15.615 | -2.261 | 0.024* | -65.907 | -4.696 |
| ERS | -3.3133 | 3.901 | -0.849 | 0.396 | -10.958 | 4.332 |
| PR | 9.7653 | 10.609 | 0.920 | 0.357 | -11.028 | 30.558 |
| QT | -4.6283 | 7.551 | -0.613 | 0.540 | -19.427 | 10.171 |
| age | 1.4154 | 0.281 | 5.031 | 0.000* | 0.864 | 1.967 |
| genders | 1.6855 | 0.579 | 2.911 | 0.004* | 0.551 | 2.821 |
| hypertension | 1.5540 | 0.515 | 3.015 | 0.003* | 0.544 | 2.564 |
| diabetes | 1.4032 | 0.490 | 2.865 | 0.004* | 0.443 | 2.363 |
| dyslipidemia | 2.8647 | 0.530 | 5.408 | 0.000* | 1.826 | 3.903 |
| smoke | 0.2107 | 0.557 | 0.378 | 0.705 | -0.881 | 1.303 |

In the previous AI-based CAD studies using CAG results as labels, the usual criterion for severe stenosis was *>* 70% lumen diameter stenosis [13, 14]. We selected 90% as the criterion for severe stenosis cases because the current guidelines specify that stenoses less than 90% are not necessarily functionally significant (i.e. they do not always induce myocardial ischaemia) [19]. The guidelines also suggest revascularization on lesions fractional flow reserve (FFR) *leq*0.8 which matches coronary stenosis ≥ 90% [20]. Therefore, predicting stenosis ≥90% can be used for identifying definite myocardial ischaemia and deciding whether to perform revascularization. Although some patients still have apparently normal ECGs when having ≥90% coronary stenosis, it is very likely that some specific changes related to ischemia have occurred that can be captured by DL models. Compared with other screening tests in medicine, such as the CHA2DS2-VASc score to assess stroke risk in patients with atrial fibrillation (0.57 0.72 AUC) [21], and brain natriuretic peptide (BNP) and N-terminal fragment of proBNP (NT-proBNP) for left ventricular dysfunction detection (AUC of 0.60 and 0.70, respectively) [22], our transfer learning DL model with ECG and baseline features achieved a higher performance with an AUC of 0.85 for distinguishing ≥ 90% coronary artery stenosis.

There are some limitations of this study. First, as discussed, the amount of data collected in this study was relatively small, which would have affected the model performance. Larger amounts of data should be used in prospective studies to externally validate model performance and verify the effects of baseline features and transfer learning. Second, the CAG results involved in this study do not have sufficient details of coronary lesions, therefore, we could only determine the severity of the stenosis and not the location of the lesion and other relevant information. Finally, although visualization tools such as Gradient-weighted Class Activation Mapping (Grad-CAM) [23] provide intuitive ways to display the feature importance for CNNs, AI ECG models are still considered to be “black-boxes” that lack interpretability, especially professional medical explanations. In Figure 6, we show the Grad-CAM mapped ECG examples that are related to the second to last layer of the transfer learning model: A is a TN example; B is an FP example; C is an FN example; D is a TP example. For the control groups A and B, the model paid more attention around the T-P segment, while for the case groups C and D, redder areas appeared around the QRS complex. However, we need to explore more nuanced approaches relevant to medical interpretations.

**Figure 6:**
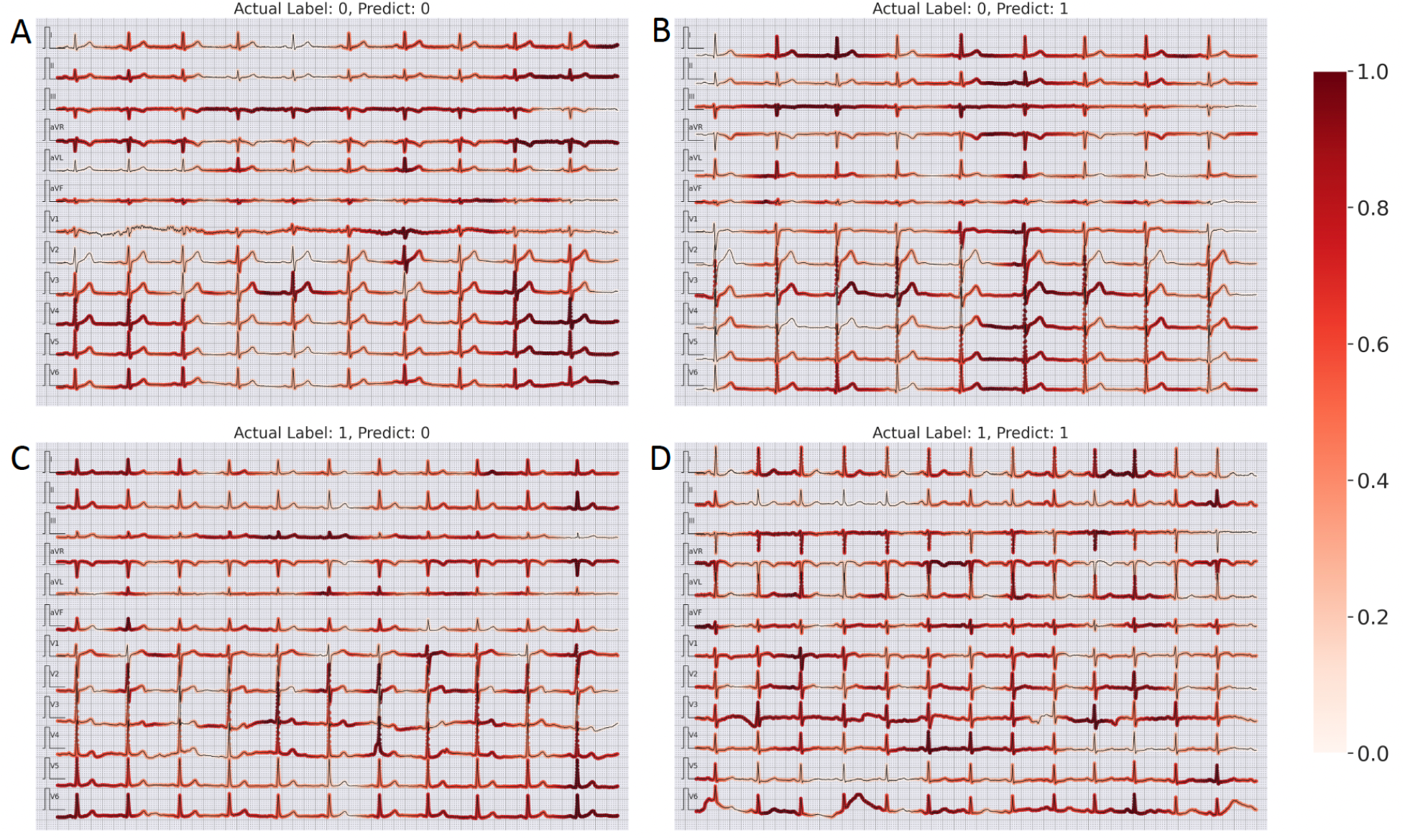
Examples of Gradcam results

## 5 Conclusion

Using apparently normal ECGs, we investigated the possibility of using DL technologies to distinguish severe coronary stenosis(≥ 90%) from no or mild coronary stenosis (< 50%). The manual readings of the selected ECGs do not show much evidence of the coronary stenosis status. Using a deep CNN trained completely with our labeled data, we could reach an accuracy of 0.683, which is higher than that achieved by cardiologists; however, the sensitivity was low. By combining the pre-trained ECG classification model parameters with patients’ baseline characteristics, we could achieve an accuracy of 0.750 with a sensitivity of 0.848 and specificity of 0.704. The findings are profound since they not only demonstrate the effectiveness of using AI ECG models to identify severe coronary stenosis, but also validate an effective approach using existing ECG models. With the existing ECG models, we can easily extract “deep features” that summarize the ECG’s inherent information and combine these features with traditional clinical data to obtain valuable auxiliary diagnostic conclusions with inexpensive calculations. Our goal is to further optimize the model to achieve early accurate identification and early intervention in high-risk patients with severe stenosis to reduce mortality and morbidity. At the same time, we hope to improve the interpretability of the model so that medical professionals have more trust in it and promote its application.

## Data Availability

Data used for this study are not publicly available due to privacy concerns. Part of the data is available upon request to the corresponding author.

## Code availability

The model code is available at https://github.com/hsd1503/resnet1d.

## Consent

All data involved in this study are obtained with the informed consent of patients and approved by the ethics committee of the Second Hospital of Tianjin Medical University (Tianjin, China).

## Funding

This work was supported by the National Natural Science Foundation of China (No. 62102008), the Research Foundation of Major Science and Technology Projects of Tianjin Municipal Science and Technology Bureau (No. 18ZXRHSY00180), the Tianjin Municipal Natural Science Foundation (No. 21JCZDJC01080), and Tianjin Key Medical Discipline (Specialty) Construction Project (No. TJYXZDXK-029A).

## Notes

### Competing Interest Statement

The authors have declared no competing interest.

### Funding Statement

This study was funded by the National Natural Science Foundation of China (No. 62102008), the Research Foundation of Major Science and Technology Projects of Tianjin Municipal Science and Technology Bureau (No. 18ZXRHSY00180), the Tianjin Municipal Natural Science Foundation (No. 21JCZDJC01080), and Tianjin Key Medical Discipline (Specialty) Construction Project (No. TJYXZDXK-029A).

### Author Declarations

Ethics committee/IRB of the Second Hospital of Tianjin Medical University (Tianjin, China) gave ethical approval for this work.

